# An Ecological Study to Investigate Links Between Atmospheric Pollutants From Farming and SARS-CoV-2 Mortality

**DOI:** 10.1101/2021.04.12.21254610

**Authors:** Paolo Contiero, Alessandro Borgini, Martina Bertoldi, Anna Abita, Giuseppe Cuffari, Paola Tomao, Maria Concetta D’Ovidio, Stefano Reale, Silvia Scibetta, Giovanna Tagliabue, Roberto Boffi, Vittorio Krogh, Fabio Tramuto, Carmelo Massimo Maida, Walter Mazzucco, “SARS-CoV-2 and Environment Working Group”

**Affiliations:** Environmental Epidemiology Unit, Fondazione IRCCS Istituto Nazionale dei Tumori, 20133 Milan, Italy; UOC Qualità dell’Aria, ARPA Sicilia, 90146 Palermo, Italy; Reporting Ambientale, Salute e Ambiente, ARPA Sicilia; 90146 Palermo, Italy; Inail-Dipartimento di medicina, epidemiologia, igiene del lavoro ed ambientale, 00078 Monte Porzio Catone (Roma), Italy; Laboratorio Tecnologie Diagnostiche Innovative Area Biologia Molecolare, Istituto Zooprofilattico Sperimentale della Sicilia, Via Rocco Dicillo 3, 90129 Palermo, Italy; Cancer Registry Unit, Fondazione IRCCS Istituto Nazionale dei Tumori, 20133 Milan, Italy; Respiratory Disease Unit, Fondazione IRCCS Istituto Nazionale Tumori, 20133 Milan, Italy; Epidemiology and Prevention Unit, Fondazione IRCCS Istituto Nazionale dei Tumori, 20133 Milan, Italy; Department of Health Promotion, Mother and Child Care, Internal Medicine and Medical Specialties (PROMISE) “G. D’Alessandro” – University of Palermo, 90127 Palermo, Italy; Regional Reference Laboratory of West Sicily for the Emergency of COVID-19, Clinical Epidemiology Unit, University Hospital “Paolo Giaccone”, 90127 Palermo, Italy; Division of Biostatistics and Epidemiology, Cincinnati Children’s Hospital Medical Centre, Cincinnati, Ohio 45229, USA; International Society of Doctors for Environment (ISDE), 52100 Arezzo, Italy; Biomarkers Unit, Fondazione IRCCS Istituto Nazionale dei Tumori, 20133 Milan, Italy

**Keywords:** particulate matter, SARS-CoV-2, mortality, pollution, ammonia, methane, nitrous oxide, farming, livestock

## Abstract

Exposure to atmospheric particulate matter and nitrogen dioxide has been linked to SARS-CoV-2 infection and death. We hypothesized that an interaction between SARS-CoV-2 infection and exposure to farming-related atmospheric pollutants worsens the effect of SARS-CoV-2 on mortality. Our objective was investigate this hypothesis by performing an ecological study in five Italian Regions (Piedmont, Lombardy, Veneto, Emilia-Romagna and Sicily) linking all-cause mortality, by province (administrative entities within regions), to atmospheric particulate matter (PM_2.5_ and PM_10_) nitrous oxide (N_2_O), ammonia (NH_3_) and methane (CH_4_) mainly produced by agricultural activities. Study outcome was change in all-cause mortality during March-April 2020, compared to March-April 2015-2019 (period) as assessed by mortality rate ratios (MRRs) estimated using multivariate negative binomial regression models that adjusted for air temperature, humidity and population density. The MRR for the interaction of period with NH_3_ exposure, considering all pollutants together was 1.133, equivalent to a 13.3% increase in mortality over and above that due to period (proxy for COVID-19 mortality) for each ton/km^2^ increase in NH_3_ emissions. Although the study was ecological, and did not provide evidence of a causal link between SARS-CoV-2 and farming-related pollutants, in accord with the precautionary principle we recommend application of measures to limit NH_3_ exposure particularly while the COVID-19 pandemic continues.

## 1. Introduction

According to the World Health Organization, on March 31, 2021, worldwide there were 128,540,982 confirmed cases of SARS-CoV-2 infection (COVID-19) and 2,808,308 deaths (2.18% of confirmed cases) [1]. The corresponding figures for Italy were 3,584,899 confirmed cases and 109,346 (3.5%) deaths [1]. These are likely to be underestimates [2].

Several studies have investigated effects of environmental and meteorological factors on the dissemination and severity of viral respiratory infections [3-6]. A 2018 paper provided evidence that air pollution from coal burning had exacerbated the “Spanish flu” pandemic of 1918 [7]. Although most COVID-19 patients develop mild or no symptoms, more severe and life-threatening symptoms such as pneumonia (often leading to acute respiratory distress syndrome), vascular inflammation and thrombosis, myocarditis, and cardiac arrhythmia, also occur, typically associated with excessive inflammation and cytokine storm [8].

Long-term and short-term exposure to atmospheric particulate matter ≤10 μm (PM_10_) and ≤2.5 μm (PM_2.5_) have been linked to respiratory and cardiovascular events [9]. A recent US study found that an increase of just 1 μg/m^3^ in long-term exposure to PM_2.5_ was associated with an 8% increase in the COVID-19 death rate [10]. Studies in Italy have identified associations between COVID-19 and COVID-19-related death and exposure to PM_10_ and PM_2.5_ [11-14]; while tropospheric nitrogen dioxide (NO2) in northern Italy was associated with levels of SARS-CoV-2 infection [15]. The COVID-19 pandemic in Italy started in the Po Valley of northern Italy – one of the most polluted areas in the world [16] – where intensive livestock rearing and heavy use of fertilizers make major contributions to atmospheric pollution [17]. In December 2019 in the Po Valley Regions of Lombardy, Piedmont, Emilia-Romagna and Veneto, there were, respectively, 1,543,639, 824,801, 627,627 and 824,112 cattle, and 3,984,633, 1,121,723, 1,377,527, and 717,557 pigs. In comparison, the island Region of Sicily had 387,619 cattle and 45,152 pigs [18]. As regards nitrogen fertilizer use, the 2019 figures (tonne/km^2^/year) were 12.92 in Lombardy, 8.27 in Piedmont, 18.64 in Veneto, and 19.50 in Emilia-Romagna, compared to 5.20 in Sicily [18].

Agricultural activity produces NH_3_, CH_4_ and N_2_O, and also PM_10_ and PM_2.5_ [19-21]. In Italy, agriculture is the main source of NH_3_ emissions, with an estimated 362.18 kilotonnes/year, accounting for 94.3% of the total [19]. In Lombardy in 2017 (latest year for which figures are available) the proportion of NH_3_ emissions from agriculture varied by province (68-99%), as did the proportions of CH_4_ and N_2_O emissions (7-87% and 27-94%, respectively) [22].

Analysis of all-cause mortality data indicates that COVID-19-related deaths are underestimated [2;23]. A study that analysed 22 countries reported that in Italy COVID-19-related deaths were underestimated by 30% with similar underestimates in several other countries [24]. For this reason all-cause mortality is the epidemiologic indicator of choice for assessing the mortality impact of COVID-19 [2;25-26] from which excess deaths can be estimated as proxy of COVID-19 mortality [25].

We hypothesized that long-term exposure to farming-related air pollutants might predispose to increased risk of COVID-19-related death. To do this we performed the preliminary investigation presented in this work: this was an ecological study, firstly to find out whether the results justified additional analytic studies. Secondly because there was a possibility of identifying modifiable risk factors to limit the effect of the COVID-19 on mortality.

## 2. Materials and Methods

### 2.1. Study Design

We conducted an ecological study to investigate whether interaction between SARS-CoV-2 infection and exposure to farming-related air pollutants worsened the effect of SARS-CoV-2 on mortality at the level of the Provinces (administrative entities within Regions) in the Italian Regions of Lombardy, Emilia-Romagna, Piedmont, Veneto, and Sicily. We investigated the period of the first COVID-19 wave (March-April 2020) in comparison with the same two-month period in the years 2015 to 2019. Mortality differences between these two periods are a good proxy for mortality due to COVID-19.

### 2.2. Pollutant Exposure

We assessed atmospheric PM_2.5_, PM_10_, N_2_O, NH_3_, CH_4_ and NO_2_. The first five were included because they are linked to farming; NO_2_ was included because it could be a confounder or effect modifier. Atmospheric PM_2.5_, PM_10_ and NO_2_ levels are measured by monitoring stations and the data archived by Regional Environmental Protection Agencies (ARPAs) [27-31]. We accessed ARPA data from the monitoring stations in the chief towns of each Province and used them as proxies for exposure of the entire provincial populations. We averaged daily concentrations of over 2019 to arrive at estimates of long-term exposure. However for PM_10_ and NO_2_ we had use the latest available data (2015) for one province, and used the data from an adjacent province in another province. We did not use 2020 data, as they could have biased estimates because of changes in pollutant levels caused by lockdown [32;33].

NH_3_ concentrations are monitored by too few stations to be representative of the areas being studied; and levels of CH_4_ and N_2_O are not measured routinely. We therefore estimated levels of atmospheric NH_3_, CH_4_ and N_2_O, again at the level of the provinces. These estimates were derived from estimates produced by the ARPAs [22;31].

ARPAs estimate NH_3_ emissions for each province and for each known source (e.g. pigs) by multiplying the average number pigs per year present in the province by an estimate of the average production by each animal (as kg NH_3_/animal/year). A similar method is applied to each type of livestock present, and also other NH_3_ sources (e.g. fertilizer use). Total NH_3_ emissions are obtained by summing estimate emissions from all known sources. Similar methods are used to estimate emissions of CH_4_ and N_2_O.

We next derived provincial exposure indices from the ARPA estimates by dividing the total estimated quantities of emissions (tonne/year) by the area (km^2^) of the province, thereby taking account of variations in province area. We used data for the latest available year as proxy for long-term exposure: 2017 for Lombardy and Emilia-Romagna (except for one province for which data were for 2016), 2015 for Piedmont and Veneto, and 2012 for Sicily.

Backes et al. [34] showed that the atmospheric concentration patterns of NH_3_ are in line with NH_3_ emission patterns, indicating the provincial exposure indices are acceptable indicators of atmospheric NH_3_. To further investigate provincial exposure indices as indicators of atmospheric pollution levels we assessed correlations (Pearson’s r) between NH_3_ emissions and cattle numbers, pig numbers and fertilizer use per km^2^ by region. We could only compare data at the regional level because of the unavailability of data on numbers of pigs and cattle and nitrogen fertilizer use at the provincial level,

### 2.3. Meteorological Variables

We hypothesized that temperature influenced COVID-19 rates over the short-term and so might have an effect on mortality. We therefore used temperature and humidity, as measured by ARPA monitoring stations in the main town of each province, as proxies for the average temperature and average relative humidity in each province, in March and April in 2020, and also in March-April 2019.

### 2.4. Additional covariates

Educational level is a potential risk factor for SARS-CoV2 infection and death [35]. We therefore downloaded provincial data on educational level from ISTAT [36] (as proportion of people who continued schooling after 14 years of age) and added these as covariates to the models. Social deprivation is also linked to SARS-CoV-2 infection and death [37]. However social deprivation indices at the provincial level were not available so we assumed that educational level was an approximate proxy for social deprivation. Income is also a component of social deprivation but was not available at provincial level so we accessed regional level data from ISTAT [40] and assessed correlations (Pearson’s r) with NH_3_ by regions to provide indications as to the possible effects of income on our findings.

High body mass index (BMI) and diabetes are recognized as major risk factors for SARS-CoV2 death [38;39] and could bias estimates of links between farming-related pollutants and SARS-CoV-2 mortality. Provincial level data were unavailable, so we accessed regional level data from ISTAT [40] and assessed correlations (Pearson’s r) with NH_3_ to provide indications as to the possible effects of BMI and diabetes on our findings.

Variation in lockdown periods across provinces could also bias the results of a study like this. However in Italy during the first COVID-19 wave, days of lockdown were closely similar throughout the country. Similarly, regulations to limit contagion (mask-wearing, social distancing) applied to the whole country during the two-month study period.

### 2.5. Outcome

All-cause daily mortality data, provided by the Italian National Statistics Institute (ISTAT) [41], was primary study outcome. Mortality data are available by municipality (administrative entities within provinces). We calculated total mortality by province, for March-April 2020, as the sum of the daily death counts in all the municipalities in each province. We also estimated average mortality in March-April of 2015-2019, as the average, over the five years, of the sum of daily death counts in all the municipalities of each province.

### 2.6. Statistical Methods

We first sought correlations (Pearson’s r) between levels of the atmospheric pollutants being studied (PM_2.5_, PM_10_, N_2_O, NH_3_, CH_4_ and NO_2_). We next analyzed associations between changes in total mortality from March-April 2015-2019 to 2020 and atmospheric pollutant levels, humidity, temperature, and population density (study covariates). To do this we used negative binomial regression models, that included a population size offset, and which estimated mortality rate ratios (MRRs) in relation to study covariates. MRR is the ratio of the mortality rate for a specific value of a covariate relative to reference. Thus, for the covariate “period” MRR is mortality in March-April 2020 relative to mortality in March-April in 2015-2019, and is a good proxy for the increase in mortality due to COVID-19. We were particularly interested in MRRs for the interaction between period and the other covariates (mainly pollutant levels) as these are an estimate of the mortality associated with the pollutant over and above that due to COVID-19 alone.

We first ran basic models that included period, one covariate, and an interaction term between period and covariate. We next ran complete models that included all covariates, and all interaction terms between period and covariates.

Information on PM_10_ levels was available for all provinces, while PM_2.5_ data were unavailable for six provinces. We therefore ran two sets of models, one that included all 45 provinces but excluded PM_2.5_, and another that included PM_2.5_ and the 39 provinces for which PM_2.5_ was available. The R statistical package [42], version 4.0.2 was used to perform the analyses.

## 3. Results

Supplementary Table S1 lists the provinces included in the study, together with their populations, population densities. Compared to mortality in March-April 2015-2019, mortality in March-April 2020 varied from a 1.8% decrease in Agrigento Province (Sicily) to a 364.3% increase in Bergamo (Lombardy). Mean values of atmospheric PM_10_, PM_2.5_ and NO_2_ across provinces were 26.01, 17.58 and 25.23 μg/m^3^ respectively; the PM_10_ ranged from 14.0 (Verbano-Cusio-Ossola, Piedmont) to 35.2 μg/m^3^ (Cremona, Lombardy), the PM_2.5_ ranged from 8.0 (Enna, Sicily) to 26.1 μg/m^3^ (Cremona, Lombardy), and the NO_2_ ranged from 4.0 (Agrigento, Sicily) to 45.7 μg/m^3^ (Monza Brianza, Lombardy).

As regards emissions, mean across-province exposure index estimates for NH_3_, CH_4_ and N_2_O were 2.22, 9.76, and 0.36 tonne/km^2^/year, respectively. The NH_3_ exposure index ranged from 0.12 (Verbano-Cusio-Ossola, Piedmont) to 10.3 tonne/km^2^/year (Cremona, Lombardy); the CH_4_ index ranged from 1.47 (Sondrio, Lombardy) to 33.17 tonne/km^2^/year (Milan, Lombardy), and the N_2_O index ranged from 0.06 (Verbano-Cusio-Ossola, Piedmont) to 1.12 tonne/km^2^/year (Cremona, Lombardy).

Table 1 shows Pearson correlation coefficients for atmospheric pollutants PM_10_, PM_2.5_, NO_2_, NH_3_, CH_4,_ and N_2_O.

**Table 1.**
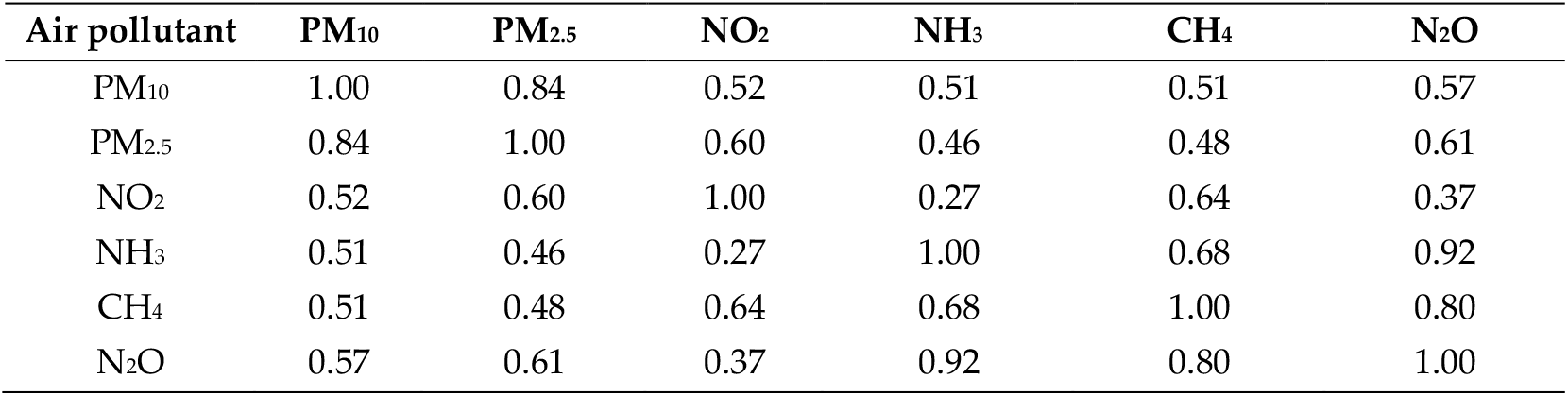
Pearson’s correlation coefficients between atmospheric pollutants

| Air pollutant | PM<sub>10</sub> | PM<sub>2.5</sub> | NO<sub>2</sub> | NH<sub>3</sub> | CH<sub>4</sub> | N<sub>2</sub>O |
| --- | --- | --- | --- | --- | --- | --- |
| PM<sub>10</sub> | 1.00 | 0.84 | 0.52 | 0.51 | 0.51 | 0.57 |
| PM<sub>2.5</sub> | 0.84 | 1.00 | 0.60 | 0.46 | 0.48 | 0.61 |
| NO<sub>2</sub> | 0.52 | 0.60 | 1.00 | 0.27 | 0.64 | 0.37 |
| NH<sub>3</sub> | 0.51 | 0.46 | 0.27 | 1.00 | 0.68 | 0.92 |
| CH<sub>4</sub> | 0.51 | 0.48 | 0.64 | 0.68 | 1.00 | 0.80 |
| N<sub>2</sub>O | 0.57 | 0.61 | 0.37 | 0.92 | 0.80 | 1.00 |

Table 2 presents modelling results for all 45 provinces included in the study: the MRRs represent percentage increases in overall death rate in relation to increments of 1 μg/m^3^ in PM_10_, and NO_2_, 1 tonne/km^2^ increments in NH_3_, CH_4_ and N_2_O, a 1°C increment in atmospheric temperature, a 1% increment in relative humidity, a one unit increase in population density, or in people who continued schooling after 14 years of age compared to those who attended only up to 14 years.

**Table 2.** Mortality rate ratios (MRRs)* with 95% confidence intervals (CI) for all 45 Italian provinces included in the study.

| Variable | MRR* (95%CI) |  |
| --- | --- | --- |
|  | Basic model<sup>§</sup> | Complete model<sup>‡</sup> |
| PM<sub>10</sub> | 0.997 (0.981-1.013) | 1.003 (0.985-1.021) |
| PM<sub>10</sub> and period interaction | 1.027 (1.004–1.050) | 0.995 (0.970–1.020) |
| NO<sub>2</sub> | 0.996 (0.986-1.006) | 1.000 (0.987-1.013) |
| NO<sub>2</sub> and period interaction | 1.025 (1.010-1.040) | 1.017 (0.999-1.037) |
| NH<sub>3</sub> | 0.992 (0.958-1.029) | 0.997 (0.909-1.088) |
| NH<sub>3</sub> and period interaction | 1.094 (1.040-1.151) | 1.133 (1.003–1.281) |
| N<sub>2</sub>O | 0.882 (0.623-1.266) | 0.721 (0.249-2.91) |
| N<sub>2</sub>O and period interaction | 2.195 (1.340-3.595) | 0.243 (0.056-1.128) |
| CH<sub>4</sub> | 0.994 (0.982-1.007) | 1.012 (0.984-1.041) |
| CH<sub>4</sub> and period interaction | 1.032 (1.015-1.051) | 1.043 (1.001-1.087) |
| Temperature | 0.983 (0.901-1.072) | 0.996 (0.928-1.070) |
| Temperature and period interaction | 0.961 (0.839-1.102) | 0.980 (0.881–1.090) |
| Humidity | 1.003 (0.990 -1.016) | 1.002 (0.989-1.015) |
| Humidity and period interaction | 0.995 (0.978-1.012) | 0.987 (0.970-1.003) |
| Population density | 1.000 (0.999-1.001) | 1.000 (0.999-1.001) |
| Population density and period interaction | 1.000 (0.999-1.001) | 1.000 (0.999-1.001) |
| Education | 0.991 (0.965-1.019) | 1.004 (0.973-1.035) |
| Education and period interaction | 1.022 (0.983-1.063) | 0.992 (0.950-1.036) |
| Period (2020 vs. 2015-2019) | 1.739 (1.526-1.983) | 4.850 (0.30-78.881) |

\* MMRs are percentage increases in overall death rate associated with: 1 $\mu\text{g}/\text{m}^3$ increase in atmospheric $\text{PM}_{10}$ ; 1 $\mu\text{g}/\text{m}^3$ increase in atmospheric $\text{NO}_2$ ; 1 tonne/year/ $\text{km}^2$ increases in emissions of $\text{NH}_3$ , $\text{CH}_4$ or $\text{N}_2\text{O}$ ; 1°C increase in atmospheric temperature, 1% increase in relative humidity, one unit increase in population density, or in people who continued schooling after 14 years of age compared to those who attended only up to 14 years.
§ Basic models include only period, a single covariate and its interaction with period.
‡ Complete models include all covariates together with their interactions with period.

The basic regression models identified significantly increased MMRs for interactions between period and: PM_10_ (MRR: 1.027; 95%CI: 1.004–1.050),NO_2_ (MRR: 1.025; 95%CI: 1.010-1.040), NH_3_ (MRR: 1.094; 95%CI: 1.040-1.151), N_2_O (MRR: 2.195; 95%CI: 1.340-3.595) and CH_4_ (MRR: 1.032; 95%CI: 1.015-1.051) indicating that high levels of these pollutants were significantly linked to an increase in total mortality over and above that due to period (proxy for presence of COVID-19). None of the individual covariates was significantly associated with MRR.

In the complete models, MRRs for the interaction between CH_4_ and period (MRR: 1.043; 95%CI: 1.001–1.087) and also for the interaction between NH_3_ and period (MRR: 1.133; 95%CI: 1.003–1.281) remained significant, indicating that an increase of 1 tonne/km^2^/year in NH_3_ emissions was significantly associated with a 13.3% increase in all-cause death over and above that associated with period (proxy for COVID-19). MRRs for interaction of period with PM_10_, N_2_O and NO_2_ all reduced, compared to the basic model, and became non-significant.

Table 3 shows results for the 39 provinces for which PM_2.5_ data were available. The MRRs were similar to those obtained for all 45 provinces, except that in the basic model, MRR for interaction between period and PM_2.5_ was significant, but for PM_10_ was not. As regards the complete models, only the MRR for interaction between period and NH_3_ was significant (1.145; 95%CI: 1.006–1.305) corresponding to a 14.5% increase (over and above that for period alone) in mortality for each tonne/km^2^/year increase in NH_3_ emissions.

**Table 3.**
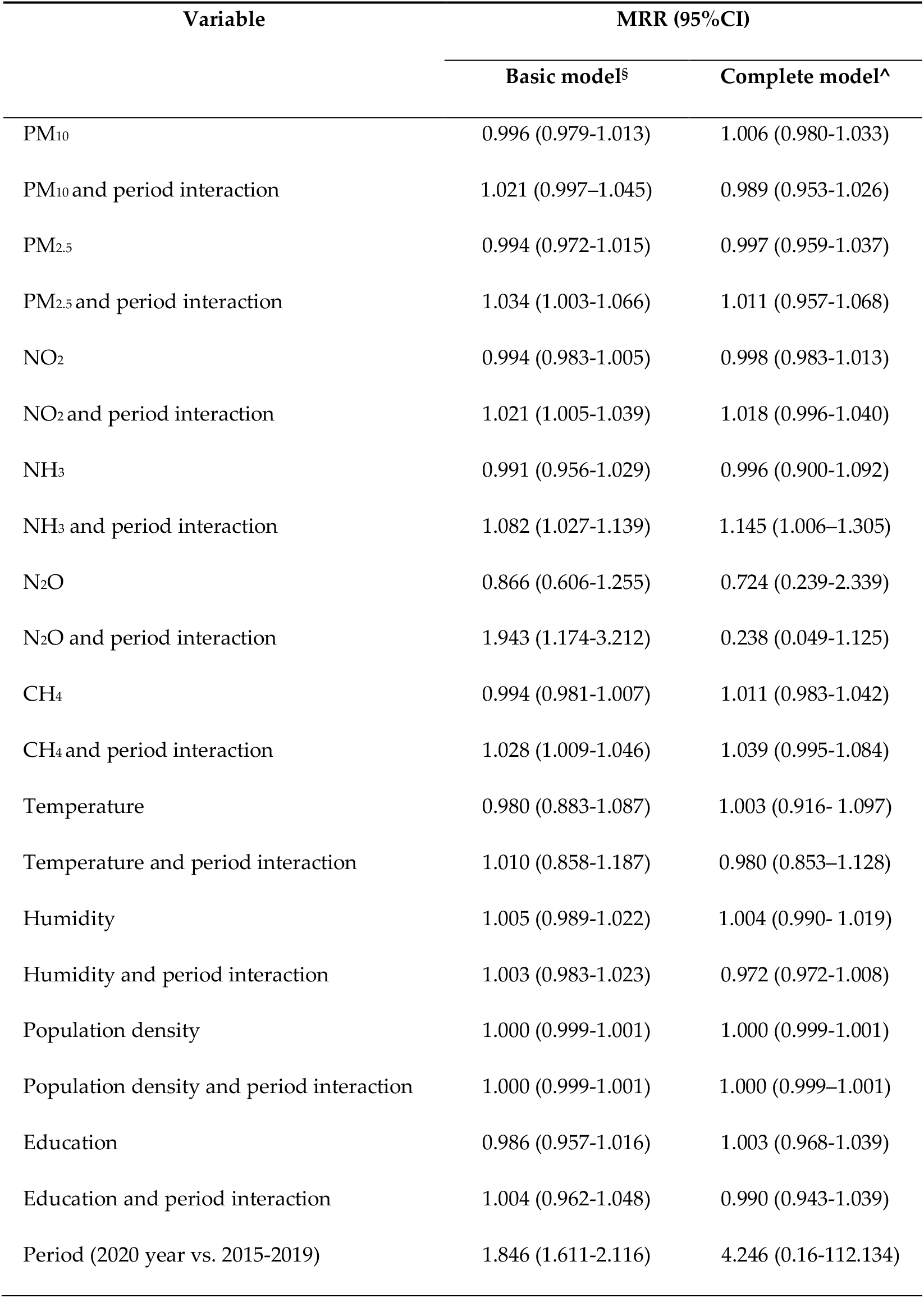
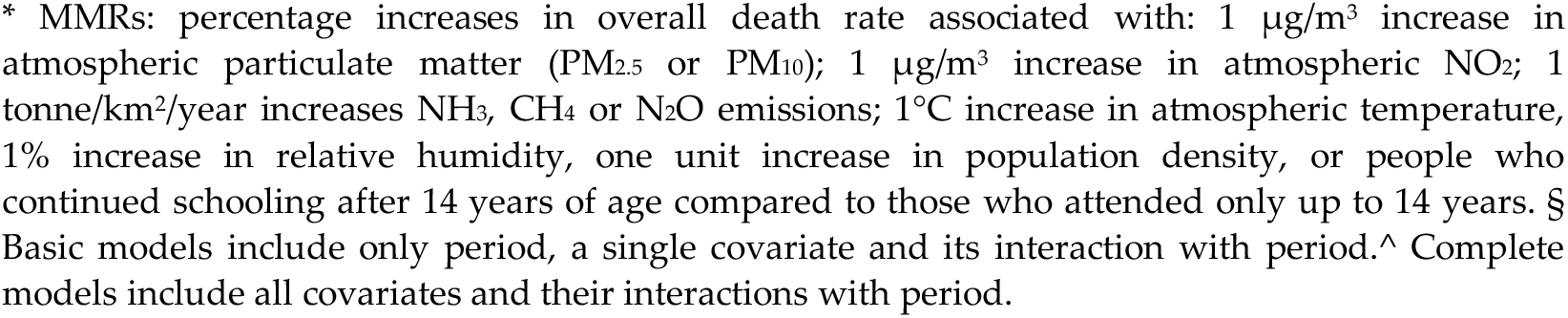
Mortality rate ratios (MRRs)* with 95% confidence intervals (CI). Analyses restricted to the 39 Italian provinces for whichPM2.5 data were available.

| Variable | MRR (95%CI) |  |
| --- | --- | --- |
|  | Basic model<sup>§</sup> | Complete model<sup>^</sup> |
| PM<sub>10</sub> | 0.996 (0.979-1.013) | 1.006 (0.980-1.033) |
| PM<sub>10</sub> and period interaction | 1.021 (0.997-1.045) | 0.989 (0.953-1.026) |
| PM<sub>2.5</sub> | 0.994 (0.972-1.015) | 0.997 (0.959-1.037) |
| PM<sub>2.5</sub> and period interaction | 1.034 (1.003-1.066) | 1.011 (0.957-1.068) |
| NO<sub>2</sub> | 0.994 (0.983-1.005) | 0.998 (0.983-1.013) |
| NO<sub>2</sub> and period interaction | 1.021 (1.005-1.039) | 1.018 (0.996-1.040) |
| NH<sub>3</sub> | 0.991 (0.956-1.029) | 0.996 (0.900-1.092) |
| NH<sub>3</sub> and period interaction | 1.082 (1.027-1.139) | 1.145 (1.006-1.305) |
| N<sub>2</sub>O | 0.866 (0.606-1.255) | 0.724 (0.239-2.339) |
| N<sub>2</sub>O and period interaction | 1.943 (1.174-3.212) | 0.238 (0.049-1.125) |
| CH<sub>4</sub> | 0.994 (0.981-1.007) | 1.011 (0.983-1.042) |
| CH<sub>4</sub> and period interaction | 1.028 (1.009-1.046) | 1.039 (0.995-1.084) |
| Temperature | 0.980 (0.883-1.087) | 1.003 (0.916- 1.097) |
| Temperature and period interaction | 1.010 (0.858-1.187) | 0.980 (0.853-1.128) |
| Humidity | 1.005 (0.989-1.022) | 1.004 (0.990- 1.019) |
| Humidity and period interaction | 1.003 (0.983-1.023) | 0.972 (0.972-1.008) |
| Population density | 1.000 (0.999-1.001) | 1.000 (0.999-1.001) |
| Population density and period interaction | 1.000 (0.999-1.001) | 1.000 (0.999-1.001) |
| Education | 0.986 (0.957-1.016) | 1.003 (0.968-1.039) |
| Education and period interaction | 1.004 (0.962-1.048) | 0.990 (0.943-1.039) |
| Period (2020 year vs. 2015-2019) | 1.846 (1.611-2.116) | 4.246 (0.16-112.134) |

\* MMRs: percentage increases in overall death rate associated with: 1 $\mu\text{g}/\text{m}^3$ increase in atmospheric particulate matter ( $\text{PM}_{2.5}$ or $\text{PM}_{10}$ ); 1 $\mu\text{g}/\text{m}^3$ increase in atmospheric $\text{NO}_2$ ; 1 tonne/ $\text{km}^2$ /year increases $\text{NH}_3$ , $\text{CH}_4$ or $\text{N}_2\text{O}$ emissions; 1°C increase in atmospheric temperature, 1% increase in relative humidity, one unit increase in population density, or people who continued schooling after 14 years of age compared to those who attended only up to 14 years. § Basic models include only period, a single covariate and its interaction with period. ^ Complete models include all covariates and their interactions with period.

Correlation analysis found the following correlations: 0.92 between provincial exposure index for NH_3_ and number of pigs per km^2^; 0.96 between provincial exposure index for NH_3_ and number of cattle per km^2^; 0.53 between the provincial exposure index for NH_3_ and number of cattle per km^2^. In addition correlations were: −0.87 between BMI and the provincial exposure index for NH_3_; and −0.77 between diabetes and the provincial exposure index for NH_3_; and 0.42 between yearly average income and the regional exposure index for NH_3_. These findings suggest that emission levels are a reasonable surrogate for atmospheric levels. Given the strength of the association of COVID-related mortality increase (13.3%) due to NH_3,_ it is unlikely that BMI, diabetes and income would have had biased this association.

## 4. Discussion

Using basic models, which excluded most covariates, we found that higher NH_3_, CH_4_, and N_2_O emissions – whose major source is agriculture – were associated with all-cause mortality increases in 2020,compared to previous years, over and above those due to period (proxy for COVID-19–related mortality). Higher PM_10_, PM_2.5_ and NO_2_ were also associated with all-cause mortality increases over and above those due to period.

Using complete models, that included all other studied pollutants as covariates, higher NH_3_ levels remained associated with a significant all-cause mortality increase: a one tonne/km^2^ increase in NH_3_ was associated with a 13.3% increase in mortality over and above that due to period. High CH_4_ levels also remained associated with a significant all-cause mortality increase in the complete model that includes all provinces, but not in the complete model that only included provinces for which for which PM_2.5_ data were available.

The fact that interactions of period with N_2_O, PM_10_, PM_2.5_ (and to some extent CH_4_) were not associated with excess mortality in the complete models is likely due to the fact that levels of these pollutants correlated highly with each other.

A major study assumption was that the provincial exposure index based on pollutant emissions is a good proxy for levels of that pollutant in the atmosphere. This assumption in supported by the study of Backes et al. [34] which showed that atmospheric concentration patterns of NH_3_ are in line with NH_3_ emission patterns. To further investigate provincial exposure indices as indicators of atmospheric pollution levels we assessed correlations (Pearson’s r) between NH_3_ emissions and cattle numbers, pig numbers and nitrogen fertilizer use, per km^2^, by region (provincial data unavailable): we found high or good correlations (0.96, 0.92 and 0.53, respectively) indicating that estimated emissions are consistent with the emission sources present, and suggesting that these estimated emissions (standardized by area size) are a reasonable proxy for atmospheric NH_3_ concentrations.

It is known that BMI and diabetes are major risk factors for COVID-19 infection and death. However provincial data for BMI and diabetes were not available and could not be included in our models as covariates. We indirectly assessed whether the association between farming-related pollutants and SARS-CoV-2 mortality were biased by BMI, diabetes and income by estimating correlations between BMI and NH_3_, between diabetes and NH_3_, and between income NH_3_ at the regional level. As noted, given the strength of the association of COVID-related mortality increase (13.3%) due to NH_3,_ it is unlikely that BMI, diabetes and income would exerted a major bias on this association. Any influence of population density and educational level on associations was taken account of by entering these as covariates in the models.

To our knowledge this is the first study to investigate relationships between farming-related atmospheric pollution and COVID-19-related mortality. However, a 2020 study conducted in Italy found that the number of SARS-CoV-2 infections per unit population depended on the type of rural landscape [43]. The study classified rural landscapes into: (A) urban and periurban with high intensity agriculture; (B) high intensity agriculture; (C) medium intensity agriculture (hilly areas), (D) low intensity agriculture (high hills and mountains). They found that areas of less intense agriculture (C, D) had fewer COVID-19 cases (per 10,000 inhabitants) than areas of intensive agriculture (A and B). Notwithstanding the markedly different methodologies, both our study and [43] identified intensive farming as a risk factor for COVID-19.

Similarly, the 2020 Lancet Countdown report highlighted a link between SARS-CoV-2 infection, environmental degradation and climate change; and identified agriculture one of the main sources of pollutant emissions [44].

Other studies have reported associations between exposure to livestock farming and respiratory diseases. Thus, a Netherlands study found that people living in high livestock density areas had a significantly higher prevalence of pneumonia than those living in low density areas [45]. A German study [46] found that people exposed to higher levels of NH_3_ had poorer pulmonary health and were more likely to be sensitized to ubiquitous allergens, than persons with lower NH_3_ exposure. A study performed in an agricultural region of Washington State (US) found that industrial-scale animal feeding operations were associated with higher daily outdoor NH_3_ levels, and that forced expiratory volume in 1 second was lower with each interquartile increase in the previous day’s NH_3_ exposure; however no associations with asthma symptoms were observed [47]. The European Academy of Allergy and Clinical Immunology position paper [48] has highlighted that increased risks of developing the respiratory diseases and allergies are associated with animal farming, and emphasized the need to protect workers from such risks.

The data provided by the above cited-studies [45-48] suggests that a link between excess COVID-19-related mortality and farming-related atmospheric pollutants is biologically plausible. The specific association we found between COVID-19-related mortality and NH_3_ might be explained by the hypothesis that atmospheric NH_3_ leads to the formation of alkaline aerosol which triggers a conformation change in the SARS-CoV-2 spike that facilitates fusion of the viral envelope with the plasma membrane of target cells [49]. It is noteworthy that clusters of SARS-CoV-2 infection have been reported in association with slaughterhouses [50] which are known to have elevated levels of NH_3_ [51].

We used change in all-cause total mortality in 2020 as a proxy for SARS-CoV-2 mortality, and to our knowledge our study is the first to do this. A strength of this approach is that it avoids loss of deaths due to underreporting of SARS-CoV-2 mortality [25]. However the pandemic may have stressed health resources so that more people than usual may have died from non-COVID-19 causes, somewhat inflating all-cause mortality. The main limitations of the study are that, for atmospheric pollutants that were measured, we used levels measured at the monitoring station in the chief town of the province as proxy for exposure of the entire provincial population. In addition, for the atmospheric pollutants NH_3_ CH_4_ and N_2_O, measured levels were not available and we used estimated emissions at the provincial level obtained from the regional environmental protection agencies, as proxies for exposure of the provincial populations.

## 5. Conclusions

Although the study was ecological, and could not identify causal links between farming-related atmospheric pollutants and COVID-19 mortality, it suggests that studies to further address this issue would be worthwhile, particularly since other data also indicate links between farming-derived pollutants and worsened outcomes of SARS-CoV-2 and other viral infections.

The agricultural sector is responsible for most NH_3_ emissions [19]. In view of the continuing severity of the SARS-CoV-2 pandemic it is important to implement measures that can speedily reduce NH_3_ emissions from agriculture, for example by covering tanks containing slurry [53]. However over the medium term structural changes are required to reduce farming-related pollution. Consumption of meat, particularly red meat, is recognized as a threat to human health and wellbeing in part because of the effect of livestock rearing on the environment, as documented the 2020 Lancet Countdown Report [44]. The desire to consume meat encourages the use of intensive farming resulting in increase emission of farming-related pollutants. As urged by the Lancet Report [44] and by Agnoletti et al. [43] we need to move towards more sustainable agriculture with reduced livestock raising. Finally we note that intensive agriculture is also a major cause of biodiversity loss worldwide, which has been implicated in the emergence of SARS-CoV-2 and other viruses [44;52].

## Data Availability

This study we used only aggregated data available on web-sites accessible to the public.

## Author Contributions

Conceptualization, P.C. and W.M.; Methodology, PC; Validation, A.A., G.C., A.B., M.B., P.T., M.C.D., S.R., S.S., G.T., R.B., V.K., F.T., C.M.M, V.R., C.C., G.L., A.C., A.R., C.V., C.DM.; Formal Analysis, P.C., V.P., M.Z.; Investigation, P.C., A.B., M.B., A.C.; Writing – Original Draft Preparation, P.C., W.M.; Writing – Review & Editing, all authors; Supervision, P.C., W.M.; Project Administration, P.C. All authors have read and agreed to the published version of the manuscript.

## Funding

This research received no external funding

## Informed Consent Statement

This study used only aggregated data available on web-sites accessible to the public.

## Data Availability Statement

This study we used only aggregated data available on web-sites accessible to the public.

## Acknowledgments

The authors thank Don Ward for help with the English and for a critical reading of the manuscript.

## Conflicts of Interest

The authors declare no conflicts of interest.

---

**Table S1.** Regions, provinces, areas, populations, included in the analysis.

| Region | Province | Area<br>(km<sup>2</sup>) | Population | Population<br>density (km<sup>-2</sup>) |
| --- | --- | --- | --- | --- |
| Piemonte | Alessandria | 3558.8 | 420017 | 118.0 |
| Piemonte | Asti | 1510.1 | 214342 | 141.9 |
| Piemonte | Biella | 913.2 | 174718 | 191.3 |
| Piemonte | Cuneo | 6905.0 | 586814 | 84.9 |
| Piemonte | Novara | 1340.2 | 369018 | 275.0 |
| Piemonte | Torino | 6827.0 | 2256108 | 330.4 |
| Piemonte | Verbano-Cusio-Ossola | 2260.9 | 157782 | 69.7 |
| Piemonte | Vercelli | 2081.6 | 170298 | 81.8 |
| Lombardy | Bergamo | 2754.9 | 1112187 | 400.0 |
| Lombardy | Brescia | 4785.6 | 1265954 | 264.5 |
| Lombardy | Como | 1279.0 | 599204 | 468.5 |
| Lombardy | Cremona | 1770.5 | 358955 | 202.7 |
| Lombardy | Lecco | 805.6 | 337380 | 418.8 |
| Lombardy | Lodi | 783.0 | 230198 | 230.2 |
| Lombardy | Monza Brianza | 405.5 | 873935 | 2159.8 |
| Lombardy | Milano | 1575.6 | 3250315 | 2028.9 |
| Lombardy | Mantova | 2341.4 | 412292 | 176.1 |
| Lombardy | Pavia | 2968.6 | 545888 | 183.9 |
| Lombardy | Sondrio | 3195.8 | 181095 | 56.6 |
| Lombardy | Varese | 1198.1 | 890768 | 743.5 |
| Veneto | Belluno | 3610.2 | 202950 | 56.0 |
| Veneto | Padova | 2144.1 | 937908 | 438.4 |
| Veneto | Rovigo | 1819.3 | 234937 | 128.5 |
| Veneto | Treviso | 2479.8 | 887806 | 358.5 |
| Veneto | Venezia | 2472.9 | 853338 | 344.6 |
| Veneto | Verona | 3096.3 | 926497 | 300.0 |
| Veneto | Vicenza | 2722.5 | 862418 | 316.8 |
| Emilia Romagna | Bologna | 3703.0 | 1014619 | 274.7 |
| Emilia Romagna | Forlì-Cesena | 2378.4 | 394627 | 165.9 |
| Emilia Romagna | Ferrara | 2635.1 | 345691 | 131.1 |
| Emilia Romagna | Modena | 2688.0 | 705393 | 262.4 |
| Emilia Romagna | Parma | 3447.4 | 451631 | 131.1 |
| Emilia Romagna | Piacenza | 2585.8 | 287152 | 111.2 |
| Emilia Romagna | Ravenna | 1859.4 | 389456 | 209.3 |
| Emilia Romagna | Reggio Emilia | 2291.2 | 531891 | 232.1 |
| Emilia Romagna | Rimini | 864.8 | 339017 | 392.4 |
| Sicily | Agrigento | 3052.0 | 435137 | 142.5 |
| Sicily | Caltanissetta | 2128.0 | 271758 | 130.0 |
| Sicily | Catania | 3573.0 | 1108040 | 310.0 |
| Sicily | Enna | 2574.0 | 164855 | 64.0 |
| Sicily | Messina | 3266.0 | 627251 | 192.0 |
| Sicily | Palermo | 5009.0 | 1253356 | 250.2 |
| Sicily | Ragusa | 1623.0 | 320795 | 197.5 |
| Sicily | Siracusa | 2124.0 | 397341 | 187.1 |
| Sicily | Trapani | 2469.0 | 428192 | 173.4 |

## Notes

### Competing Interest Statement

The authors have declared no competing interest.

### Author Declarations

being an ecological epidemiological study, this study has the exemption of the ethics committee, as there are no sensitive patient data or clinical trials as there are no sensitive patient data or clinical trials

